# A Cross-Sectional Study on the Current Status and Determinants of Human Papillomavirus Infection in Women from Suzhou, China

**DOI:** 10.1101/2025.10.16.25338161

**Authors:** Haonan Cheng, Likun Wang, Zhemin Zhou, Heng Li, Yahui Zhan

## Abstract

This study investigated the human papillomavirus (HPV) infection status, genotype distribution, and associated risk factors among women in Suzhou to provide a theoretical basis for controlling cervical cancer and guiding vaccine development. From November 2022 to April 2023, 566 qualified participants undergoing health examinations at Suzhou Municipal Hospital were enrolled. Cervical exfoliated cells were tested for 21 HPV genotypes using fluorescence quantitative PCR, and data on general information, lifestyle, and clinical history were collected via questionnaire. The overall HPV infection rate was 11.31% (64/566). The most prevalent genotypes were HPV16, HPV52, and HPV58, each with an infection rate of 1.77%. Single infections predominated (8.83%, 50/566), with HPV16 (18%), HPV58 (14%), HPV51 (12%), and HPV81 (12%) being the most common. Multiple infections accounted for 2.47%, primarily dual infections (78.57%). Infection rates varied significantly by age, with the highest prevalence in women aged ≥50 years (24.37%), followed by those aged 40-49 (8.42%) and ≤39 (7.35%). Significant risk factors for HPV infection included older age (OR=1.075, P<0.01), secondhand smoke exposure (OR=2.126, P<0.05), a history of underlying diseases, and a history of cervical surgery. In conclusion, the HPV infection rate in Suzhou remains considerable, with genotypes 16, 52, and 58 being predominant and single infections most common. Older age, secondhand smoke exposure, underlying diseases, and cervical surgery history are key risk factors, highlighting the need for targeted prevention and comprehensive control strategies.

## Introduction

Cervical cancer represents a major global health challenge, ranking as the fourth most common cancer among women. Statistics from 2018 revealed approximately 568,800 new cases and 311,400 deaths worldwide annually, with developing countries bearing a disproportionate burden—accounting for about 85% of both incidence and mortality (Arbyn et al., 2020). China faces a particularly significant impact, reporting roughly 130,000 new cases each year, which constitutes nearly 25% of the global total (Li et al., 2025).

Persistent infection with human papillomavirus (HPV) is well-established as the primary causative agent of cervical cancer (de Sanjose et al., 2010). To date, more than 100 HPV genotypes have been identified, with over 40 known to infect the female reproductive tract. Based on their oncogenic potential, these genotypes are classified into three categories: high-risk types (including HPV16, 18, and 31) that are strongly associated with malignant lesions; low-risk types (such as HPV6, 11, and 81) that typically cause benign conditions like genital warts; and suspected high-risk types (including HPV26 and 66) whose carcinogenic potential requires further investigation (Ramakrishnan et al., 2015).

The epidemiology of HPV infection demonstrates considerable geographical variation. While the global infection prevalence among women is approximately 11.7%, mainland China reports a higher rate of about 16.14% (Li et al., 2019). Regional disparities within China are also evident, with studies showing infection rates of 18% in Shenzhen, 16% in Shenyang, and 14% in Shanxi. Similarly, the distribution of predominant genotypes varies across regions, with Italy showing prevalence of HPV16, 33, and 18, compared to China where HPV16, 58, and 52 are more common (Nascimento et al., 2018).

The development of cervical cancer involves multiple factors beyond HPV infection, including early sexual debut, contraceptive use, smoking, socioeconomic status, and genetic predisposition (Braun et al., 2021). However, persistent HPV infection remains the most significant risk factor. This study therefore aimed to investigate the HPV infection status, genotype distribution, and associated risk factors among women undergoing routine health examinations in Suzhou. The findings are expected to provide valuable evidence for developing targeted prevention strategies, optimizing vaccination programs, and ultimately reducing the burden of cervical cancer in this population.

## Materials and Methods

### Study subjects

From November 2022 to April 2023, women who underwent health checkups and cervical HPV and TCT examinations at the Health Management Center of Suzhou Municipal Hospital were selected as the survey subjects for this study. A cross-sectional study was conducted. Inclusion criteria: (1) female residents living in Suzhou City; (2) voluntary participation and signing of informed consent; (3) history of sexual activity; (4) no sexual intercourse or use of vaginal medications for 48 hours. Exclusion criteria: (1) history of hysterectomy; (2) currently menstruating, pregnant, or breastfeeding; (3) receiving immunosuppressive therapy. This study was approved by the Jiangsu Provincial Medical Ethics Review Committee.

### Questionnaire

Before specimen collection, participants were given a custom questionnaire. The questionnaire and specimen were numbered and associated with each other. Professionally trained interviewers guided the participants through the questionnaire. The questionnaire included information about their ethnicity, age, education level, marital status, maternal and childbearing history, previous alcohol consumption, previous smoking history, previous HPV testing history, previous cervical cytology history, previous cervical surgery history, vaccination history, family history of cervical cancer, and contraceptive use. The interviewer explained their questions to the participants on-site. Upon receiving the questionnaire, the interviewer checked the questionnaire for compliance. Any errors or omissions were addressed, and the participants were asked to revise or supplement the information on-site. All relevant information on the participants was collected and organized.

### Cervical cytology specimen collection

The participants were instructed to assume the lithotomy position on the bed. The medical staff used a dedicated speculum to expose the cervix, using gentle techniques to avoid trauma to the cervix and vagina. First, use a sterile cotton swab to wipe away excess cervical surface secretions. Then, place a cervical brush at the cervical os and gently rotate it in one direction for 6-10 rotations, holding it for 10 seconds, to ensure sufficient cervical exfoliated cells are collected. Finally, place the brush head in cell preservation solution and label the sample with the corresponding number.

### HPV DNA extraction

To extract DNA from cervical exfoliated cells: Place 200μl of cervical exfoliated cells in a 1.5ml centrifuge tube. Add 40μl of proteinase K (20mg/ml) solution and incubate at room temperature for 15 minutes. Then, add 200μl of binding buffer and immediately vortex to mix. Incubate at 70°C for 10 minutes until the solution becomes clear. Add 100μl of isopropyl alcohol and vortex to mix. A flocculent precipitate may form. Add the solution and flocculent precipitate obtained in the previous step to an imported adsorption column AC. (Place the adsorption column in a collection tube and centrifuge at 10,000 rpm for 30 seconds. Discard the waste liquid in the collection tube. Add 500 μl of inhibitor removal solution IR and centrifuge at 12,000 rpm for 30 seconds. Discard the waste liquid. Add 700 μl of rinse buffer WB (please check whether anhydrous ethanol has been added first). Centrifuge at 12,000 rpm for 30 seconds and discard the waste liquid. Add 500 μl of rinse buffer WB and centrifuge at 12,000 rpm for 30 seconds and discard the waste liquid. Return the imported adsorption column AC to the empty collection tube and centrifuge at 13,000 rpm for 2 minutes. Remove as much rinse liquid as possible to prevent residual ethanol in the rinse liquid from inhibiting downstream reactions. Remove the imported adsorption column AC and place it in a clean centrifuge tube. Add 100 μl of elution buffer EB (preheat the elution buffer in a 65-70°C water bath) to the middle of the adsorption membrane. Incubate at room temperature for 3-5 minutes and centrifuge at 12,000 rpm. Centrifuge at 12,000 rpm for 1 minute. Add the resulting solution back to the centrifuge column, incubate at room temperature for 2 minutes, and then centrifuge at 12,000 rpm for 1 minute.

### Primer and probe design

Targeting the L1 region of the human papillomavirus genome, 21 type-specific primers and probes were designed, labeled with FAM, HEX, and ROX, respectively, for the corresponding types. Fluorescence quantitative PCR was used to detect HPV infection and 21 infectious genotypes, including high-risk types 16, 18, 26, 31, 33, 35, 39, 45, 51, 52, 53, 56, 58, 59, 66, 68, 73, and 82, and low-risk types 6, 11, and 81. The probe consists of an oligonucleotide with a reporter gene at the 5’ end and a quencher gene at the 3’ end. During PCR amplification, when the probe is intact, the quencher gene is close to the reporter gene, and the fluorescence emitted by the reporter gene separates from the quencher gene, generating a fluorescent signal. The fluorescent quantitative PCR instrument automatically plots the real-time amplification curve based on the detected fluorescent signal. Amplification curve was constructed to achieve qualitative detection of human papillomavirus at the nucleic acid level. PCR amplification test: 50°C, 5 minutes, 1 cycle; 95°C, 10 minutes, 1 cycle; 95°C, 10 seconds, 45 cycles; 58°C, 40 seconds. Based on the experimental test results, the receiver operating characteristic (ROC) curve method was used to confirm that this kit contained all HPV types.

### Data Analysis

Questionnaires were entered in pairs using EpiData 3.1 and matched, and statistical analysis was performed using SPSS 27.0. Frequencies and percentages were used to describe categorical variables, and chi-square tests were used to compare categorical variables. P < 0.05 was considered statistically significant. HPV infection status was used as the dependent variable, and variables with a P < 0.1 in the univariate analysis and continuous variables were used as independent variables. A multivariate logistic regression model was constructed to analyze influencing factors.

## Results

### Basic information of the survey subjects

Of the 615 questionnaires initially collected, 566 met the study inclusion criteria. The mean age of participants was 41.41 ± 8.61 years (range: 24-67 years). Age distribution showed that 245 participants (43.29%) were ≤39 years, 202 (35.69%) were 40-49 years, and 119 (21.02%) were ≥50 years. The participants reported no secondhand smoke exposure (90.99%, 515/566) and no underlying medical conditions including diabetes, hypertension, or heart disease (93.46%, 529/566). Reproductive history indicated that 33.22% (188/566) had one or two pregnancies, while 63.43% (359/566) had a single parity. Regarding abortion history, 78.27% (443/566) had never undergone medical abortion and 67.84% (384/566) had never undergone surgical abortion. Lifestyle factors revealed that 91.17% (516/566) did not take health supplements, 77.21% (437/566) maintained fruit consumption habits, 80.92% (458/566) were non-smokers, and 83.22% (471/566) did not consume alcohol. However, 39.40% (223/566) reported frequently staying up late. Medical history data showed that 98.23% (556/566) had never taken birth control pills, 98.76% (559/566) had never used estrogen supplements, 78.09% (442/566) had no history of vaginitis, 87.63% (496/566) had no intrauterine device (IUD), and 93.29% (528/566) were sexually active. Additionally, 99.29% (562/566) had no immunodeficiency history, 90.64% (513/566) had no cervical surgery history, and 45.58% (258/566) had previously undergone HPV testing. Among all participants, 14.13% (80/566) had received HPV vaccination. Of the 69 participants who specified their vaccine type, 23.19% (16/69) received the bivalent vaccine, 50.72% (35/69) received the quadrivalent vaccine, and 26.09% (18/69) received the nonavalent vaccine. Detailed characteristics are presented in Table 1.

**Table 1.**
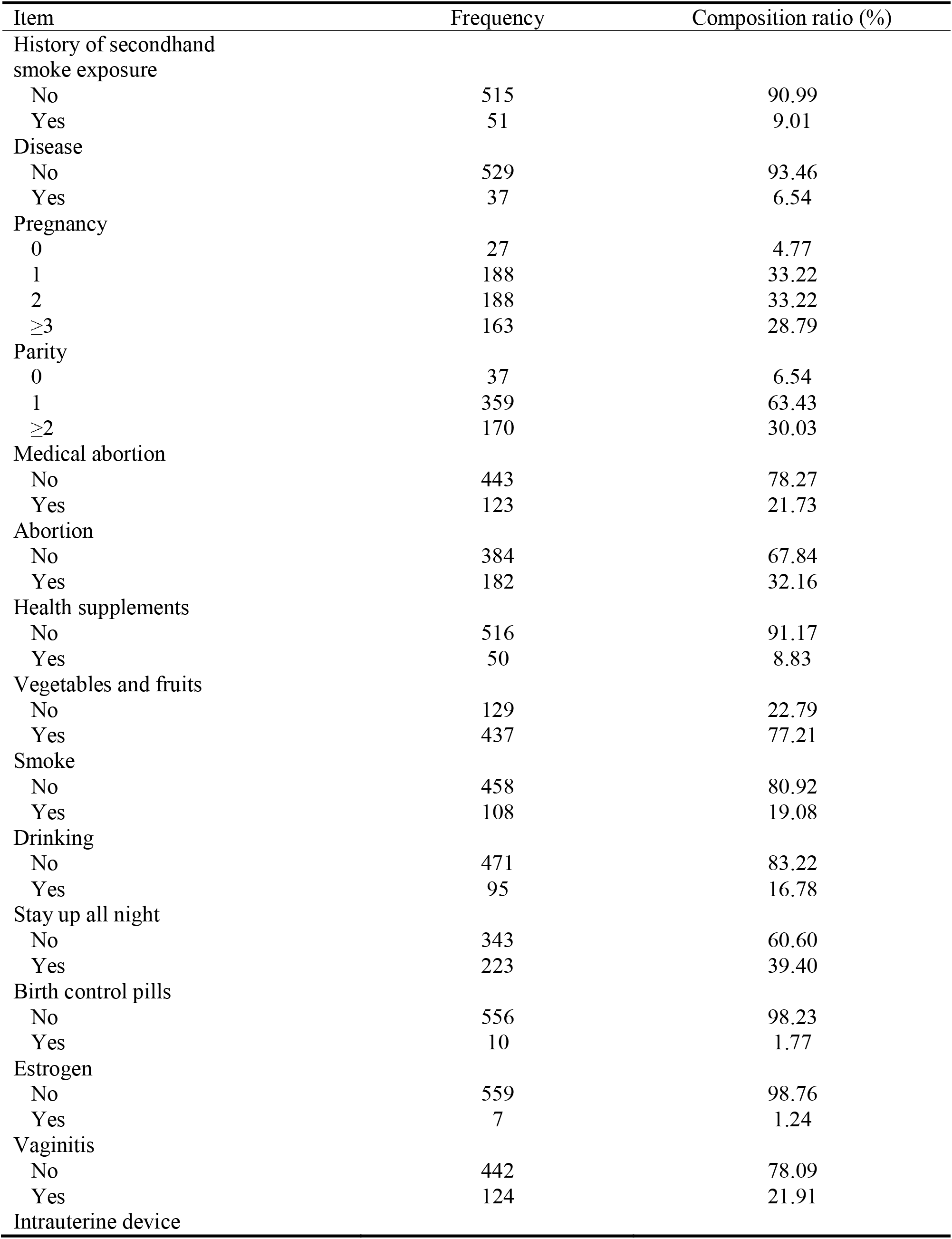
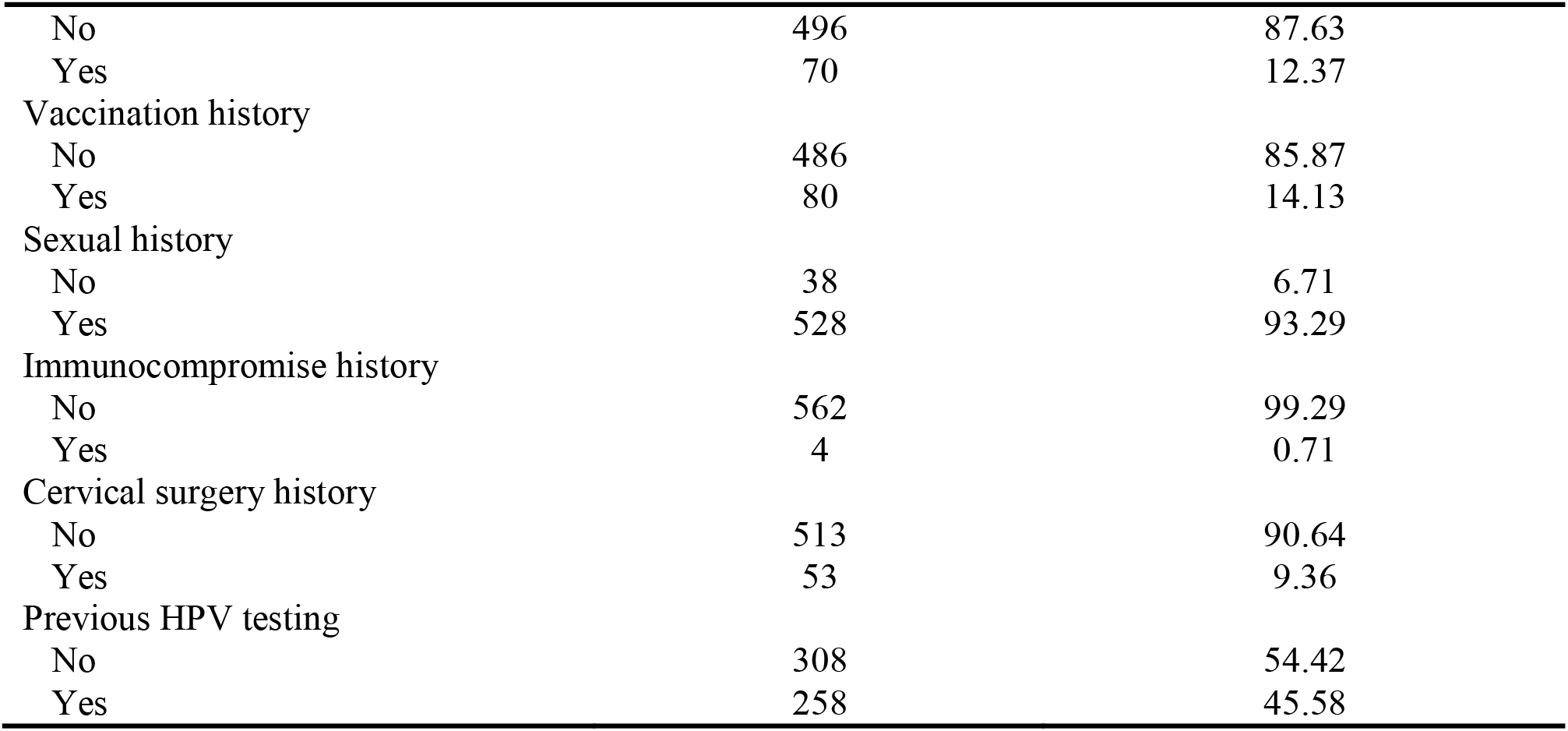
Information of the survey subjects.

| Item | Frequency | Composition ratio (%) |
| --- | --- | --- |
| History of secondhand smoke exposure |  |  |
| No | 515 | 90.99 |
| Yes | 51 | 9.01 |
| Disease |  |  |
| No | 529 | 93.46 |
| Yes | 37 | 6.54 |
| Pregnancy |  |  |
| 0 | 27 | 4.77 |
| 1 | 188 | 33.22 |
| 2 | 188 | 33.22 |
| $\geq 3$ | 163 | 28.79 |
| Parity |  |  |
| 0 | 37 | 6.54 |
| 1 | 359 | 63.43 |
| $\geq 2$ | 170 | 30.03 |
| Medical abortion |  |  |
| No | 443 | 78.27 |
| Yes | 123 | 21.73 |
| Abortion |  |  |
| No | 384 | 67.84 |
| Yes | 182 | 32.16 |
| Health supplements |  |  |
| No | 516 | 91.17 |
| Yes | 50 | 8.83 |
| Vegetables and fruits |  |  |
| No | 129 | 22.79 |
| Yes | 437 | 77.21 |
| Smoke |  |  |
| No | 458 | 80.92 |
| Yes | 108 | 19.08 |
| Drinking |  |  |
| No | 471 | 83.22 |
| Yes | 95 | 16.78 |
| Stay up all night |  |  |
| No | 343 | 60.60 |
| Yes | 223 | 39.40 |
| Birth control pills |  |  |
| No | 556 | 98.23 |
| Yes | 10 | 1.77 |
| Estrogen |  |  |
| No | 559 | 98.76 |
| Yes | 7 | 1.24 |
| Vaginitis |  |  |
| No | 442 | 78.09 |
| Yes | 124 | 21.91 |
| Intrauterine device |  |  |
| No | 496 | 87.63 |
| Yes | 70 | 12.37 |
| Vaccination history |  |  |
| No | 486 | 85.87 |
| Yes | 80 | 14.13 |
| Sexual history |  |  |
| No | 38 | 6.71 |
| Yes | 528 | 93.29 |
| Immunocompromise history |  |  |
| No | 562 | 99.29 |
| Yes | 4 | 0.71 |
| Cervical surgery history |  |  |
| No | 513 | 90.64 |
| Yes | 53 | 9.36 |
| Previous HPV testing |  |  |
| No | 308 | 54.42 |
| Yes | 258 | 45.58 |

### Regional HPV infection rate and genotype distribution

Of the 566 individuals included in the study, 64 tested positive for HPV, yielding an overall infection rate of 11.31%. The genotypes with the highest infection rates in the cohort were HPV16, HPV52, and HPV58, each with a prevalence of 1.77% (10/566). Among the HPV-positive individuals, single infections were predominant, constituting 8.83% (50/566) of the total study population. Genotype distribution within this single-infection group revealed HPV16 as the most common (18%, 9/50), followed by HPV58 (14%, 7/50), and a tie between HPV51 and HPV81 (12% each, 6/50). Other detected genotypes included HPV52 (10%, 5/50), HPV39, HPV59, and HPV31 (6% each, 3/50), HPV53, HPV66, and HPV68 (4% each, 2/50), and HPV56 and HPV82 (2% each, 1/50). Multiple infections accounted for 2.47% (14/566) of all participants. The majority of these (78.57%, 11/14) were double infections, with the combination of HPV35 and HPV52 being particularly notable. Triple or higher infections represented 21.43% (3/14) of multiple infections, specifically identified as HPV45+58+73, HPV56+66+6, and HPV39+51+59 (Figures 1).

**Figure 1.**
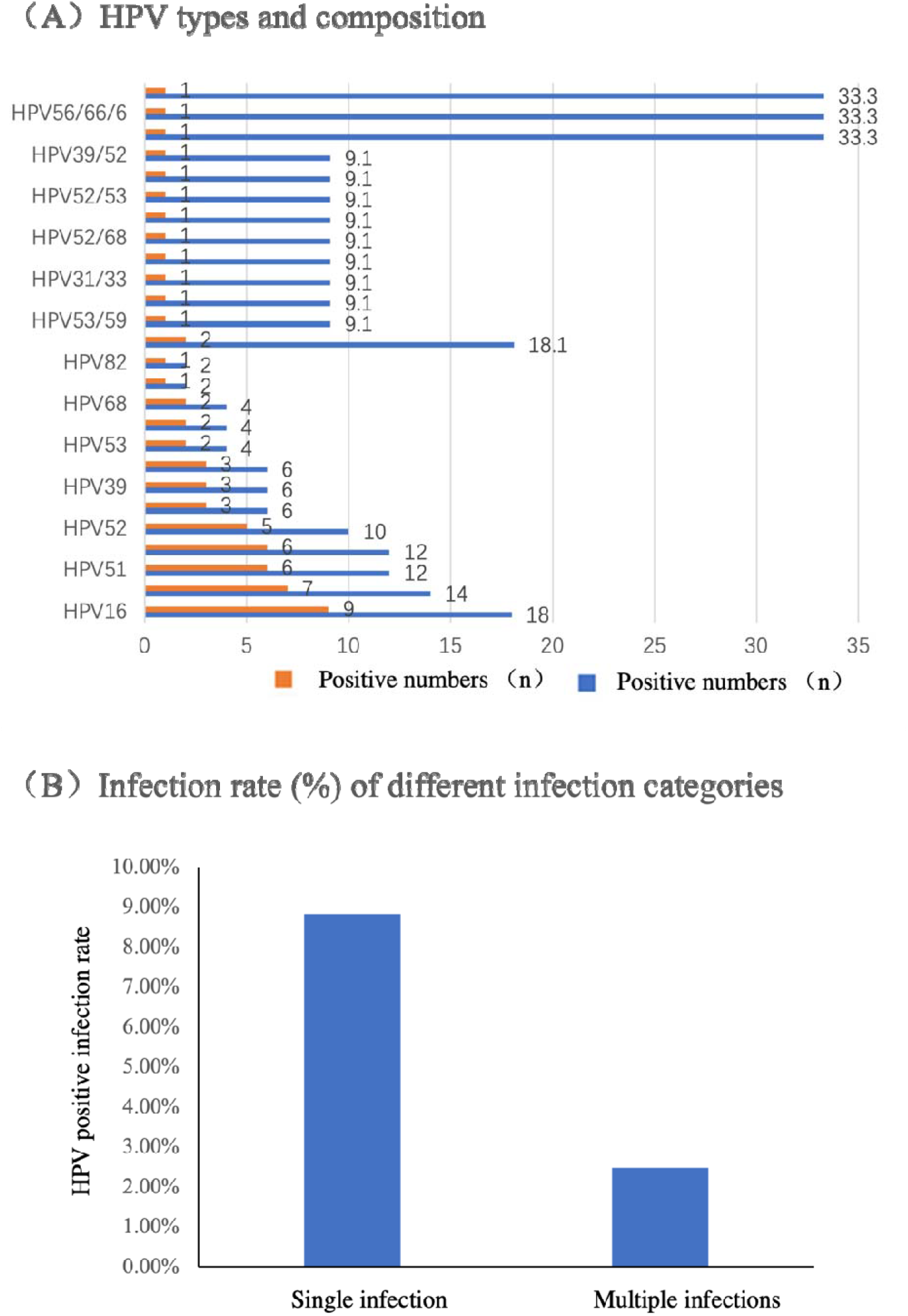
Distribution of HPV genotypes and infection patterns. (A) Frequency and composition of specific HPV genotypes detected in the study population. HPV16 (14.0%) was the most prevalent single genotype, followed by HPV51 (7.0%). Several co-infection patterns were also identified, with combinations such as HPV39/52 and HPV52/53 each accounting for 33.3% of their respective detection instances. (B) Prevalence of single versus multiple HPV infections. Single infections (8.83%) were significantly more common than multiple infections (2.47%) in the study cohort.

### Univariate analysis of factors influencing HPV infection

A significant association was observed between HPV infection rates and specific demographic and clinical characteristics (Figure 2, Table 2). The prevalence of HPV infection varied markedly across age groups (P<0.001), demonstrating an age-dependent increase. The highest infection rate was found in women aged ≥50 years (24.37%, 29/119), which was approximately three times the rate in the 40-49 age group (8.42%, 17/202) and more than three times the rate in the ≤39 age group (7.35%, 18/245). Several risk factors showed statistically significant associations with increased HPV infection rates (P<0.05). Participants with a history of secondhand smoke exposure had more than double the infection rate (21.57%, 11/51) compared to those without such exposure (10.29%, 53/515). Similarly, the presence of underlying diseases was associated with a substantially higher infection rate (24.32%, 9/37) versus those without underlying conditions (10.40%, 55/529). A history of cervical surgery also represented a significant risk factor, with infected individuals showing nearly twice the infection rate (20.75%, 11/53) compared to those without surgical history (10.33%, 53/513). These findings collectively indicate that advanced age (particularly ≥50 years), secondhand smoke exposure, underlying medical conditions, and previous cervical surgery are significant risk factors for HPV infection in this population.

**Table 2.** Univariate analysis of factors affecting HPV infection.

| Item | Frequency | Positive (n=502) | Negative (n=64) | $\chi^2$ | P |
| --- | --- | --- | --- | --- | --- |
| Age ( Year ) |  |  |  |  |  |
| ≤39 | 245 ( 43.29 ) | 227 ( 45.22 ) | 18 ( 28.13 ) | 25.762 | <0.001 |
| 40 ~ 49 | 202 ( 35.69 ) | 185 ( 36.85 ) | 17 ( 26.56 ) |  |  |
| ≥50 | 119 ( 21.02 ) | 90 ( 17.93 ) | 29 ( 45.31 ) |  |  |
| History of secondhand smoke exposure |  |  |  |  |  |
| No | 515(90.99) | 462(92.03) | 53(82.81) | 5.885 | 0.015 |
| Yes | 51(9.01) | 40(7.97) | 11(17.19) |  |  |
| Disease |  |  |  |  |  |
| No | 529(93.46) | 474(94.42) | 55(85.94) | 5.268 | 0.022 |
| Yes | 37(6.54) | 28(5.58) | 9(14.06) |  |  |
| Pregnancy |  |  |  |  |  |
| 0 | 27(4.77) | 23(4.58) | 4(6.25) | 1.072 | 0.784 |
| 1 | 188(33.22) | 170(33.86) | 18(28.12) |  |  |
| 2 | 188(33.22) | 165(32.87) | 23(35.94) |  |  |
| ≥3 | 163(28.79) | 144(28.69) | 19(29.69) |  |  |
| Parity |  |  |  |  |  |
| 0 | 37(6.54) | 33(6.57) | 4(6.25) | 0.017 | 0.992 |
| 1 | 359(63.43) | 318(63.35) | 41(64.06) |  |  |
| ≥2 | 170(30.04) | 151(30.08) | 19(29.69) |  |  |
| Medical abortion |  |  |  |  |  |
| No | 443(78.27) | 391(77.89) | 52(81.25) | 0.377 | 0.539 |
| Yes | 123(21.73) | 111(22.11) | 12(18.75) |  |  |
| Abortion |  |  |  |  |  |
| No | 384(67.84) | 344(68.53) | 40(62.50) | 0.945 | 0.331 |
| Yes | 182(32.16) | 158(31.47) | 24(37.50) |  |  |
| Health supplements |  |  |  |  |  |
| No | 516(91.17) | 460(91.63) | 56(87.50) | 1.204 | 0.272 |
| Yes | 50(8.83) | 42(8.37) | 8(12.50) |  |  |
| Vegetables and fruits |  |  |  |  |  |
| No | 129(22.79) | 114(22.71) | 15(23.44) | 0.017 | 0.896 |
| Yes | 437(77.21) | 388(77.29) | 49(76.56) |  |  |
| Smoke |  |  |  |  |  |
| No | 458(80.92) | 410(81.67) | 48(75.00) | 1.637 | 0.201 |
| Yes | 108(19.08) | 92(18.33) | 16(25.00) |  |  |
| Drinking |  |  |  |  |  |
| No | 471(83.22) | 421(83.86) | 50(78.12) | 1.339 | 0.247 |
| Yes | 95(16.78) | 81(16.14) | 14(21.88) |  |  |
| Stay up all night |  |  |  |  |  |
| No | 343(60.60) | 301(59.96) | 42(65.62) | 0.763 | 0.382 |
| Yes | 223(39.40) | 201(40.04) | 22(34.38) |  |  |
| Birth control pills |  |  |  |  |  |
| No | 556(98.23) | 492(98.01) | 64(100.00) | 0.404 | 0.525 |
| Yes | 10(1.77) | 10(1.99) | 0(0.00) |  |  |
| Estrogen |  |  |  |  |  |
| No | 559(98.76) | 497(99.00) | 62(96.88) |  | 0.182* |
| Yes | 7(1.24) | 5(1.00) | 2(3.12) |  |  |
| Vaginitis |  |  |  |  |  |
| No | 442(78.09) | 394(78.49) | 48(75.00) | 0.403 | 0.525 |
| Yes | 124(21.91) | 108(21.51) | 16(25.00) |  |  |
| Intrauterine device |  |  |  |  |  |
| No | 496(87.63) | 441(87.85) | 55(85.94) | 0.191 | 0.662 |
| Yes | 70(12.37) | 61(12.15) | 9(14.06) |  |  |
| Vaccination history |  |  |  |  |  |
| No | 486(85.87) | 428(85.26) | 58(90.62) | 1.347 | 0.246 |
| Yes | 80(14.13) | 74(14.74) | 6(9.38) |  |  |
| Sexual history |  |  |  |  |  |
| No | 38(6.71) | 37(7.37) | 1(1.56) | 2.200 | 0.138 |
| Yes | 528(93.29) | 465(92.63) | 63(98.44) |  |  |
| Immunocompromise history |  |  |  |  |  |
| No | 562(99.29) | 500(99.60) | 62(96.88) |  | 0.065* |
| Yes | 4(0.71) | 2(0.40) | 2(3.12) |  |  |
| Cervical surgery history |  |  |  |  |  |
| No | 513(90.64) | 460(91.63) | 53(82.81) | 5.204 | 0.023 |
| Yes | 53(9.36) | 42(8.37) | 11(17.19) |  |  |
| Previous HPV testing |  |  |  |  |  |
| No | 308(54.42) | 276(54.98) | 32(50.00) | 0.568 | 0.451 |
| Yes | 258(45.58) | 226(45.02) | 32(50.00) |  |  |

\*Fisher's exact probability method

**Figure 2.**
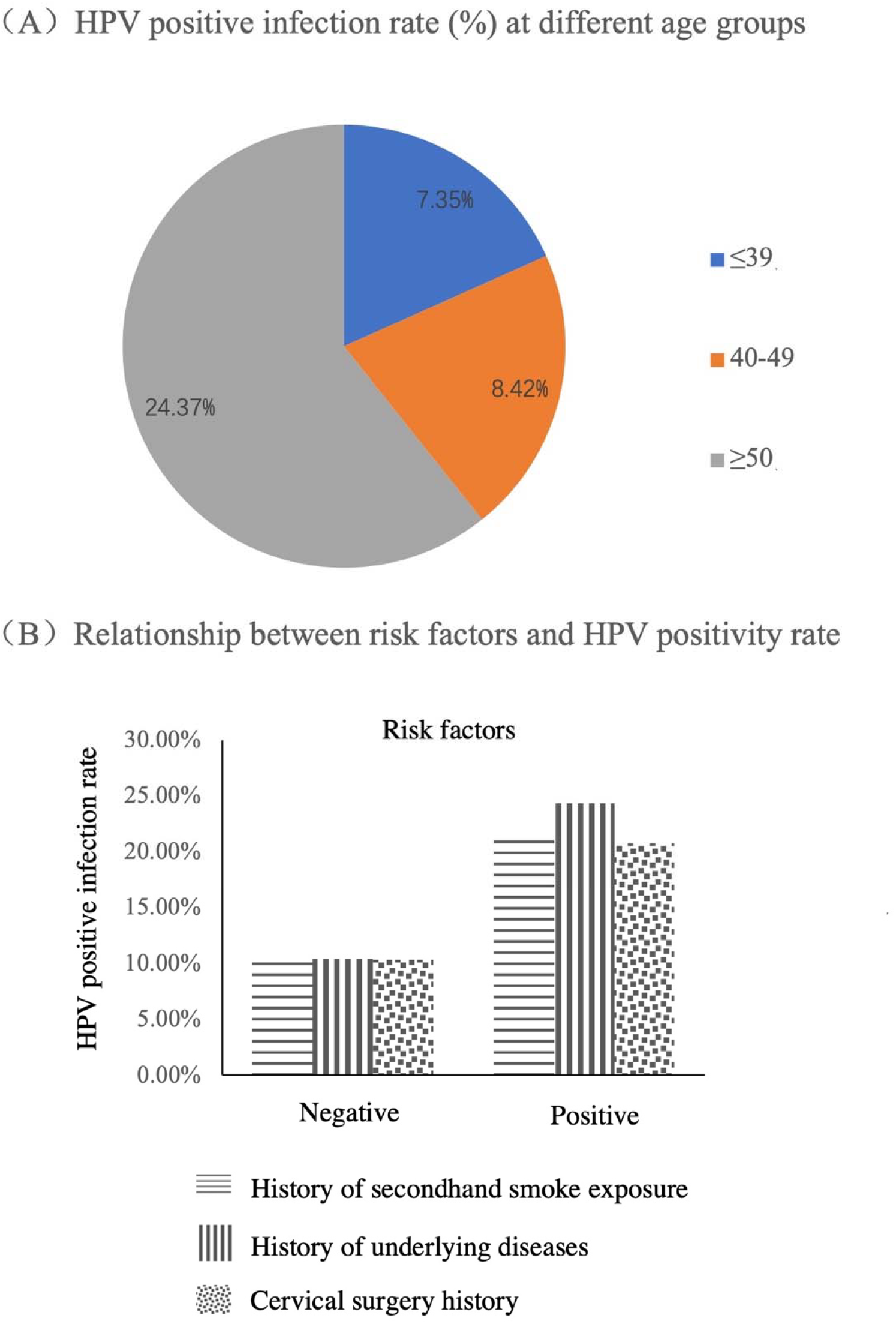
HPV infection status by age group and selected risk factors. (A) HPV positive infection rate across different age groups. The highest infection rate (24.37%) was observed in women aged ≥50 years. (B) Association between specific risk factors and HPV positivity rate. A history of underlying diseases and cervical surgery were associated with significantly higher infection rates.

### Multivariate analysis of factors affecting HPV infection

Binary logistic regression analysis was performed, incorporating factors demonstrating marginal significance (P < 0.1) in univariate analysis along with age as a clinically relevant variable. The analysis revealed that increasing age was independently associated with HPV infection risk, with each additional year of age corresponding to a 7.5% increase in infection odds (OR = 1.075, 95% CI [1.035, 1.116], P < 0.01). Furthermore, women exposed to secondhand smoke demonstrated significantly elevated risk, being 2.13 times more likely to contract HPV infection compared to their non-exposed counterparts (OR = 2.126, 95% CI [1.024, 4.415], P < 0.05). Complete regression results are presented in Table 3.

**Table 3.** Multivariate analysis of factors affecting HPV infection.

| Item | B | SE | Wald<sup>x<sup>2</sup></sup> | **P** | OR | 95% CI |
| --- | --- | --- | --- | --- | --- | --- |
| Age ( Year ) | 0.073 | 0.016 | 21.243 | <0.001 | 1.075 | 1.043-1.109 |
| History of secondhand smoke exposure |  |  |  |  |  |  |
| No * |  |  |  |  |  |  |
| Yes | 0.754 | 0.383 | 3.89 | 0.049 | 2.126 | 1.005-4.500 |

\* Control group

## Discussion

In recent years, China has witnessed a marked increase in both the incidence and mortality rates of cervical cancer, with the disease demonstrating a troubling trend toward younger onset (Wei et al., 2019). Persistent HPV infection significantly elevates the risk of developing cervical cancer. China has established a relatively comprehensive five-level cervical cancer prevention strategy, wherein primary and secondary prevention primarily rely on HPV vaccination and cervical cancer screening, respectively. Currently, three HPV vaccines are approved in China: the bivalent vaccine (targeting HPV16 and 18), the quadrivalent vaccine (targeting HPV6, 11, 16, and 18), and the nine-valent vaccine (targeting HPV6, 11, 16, 18, 31, 33, 45, 52, and 58) (Zhu et al., 2022). Investigating regional epidemiological data is crucial for understanding local HPV infection risk factors, thereby informing vaccine development and prevention strategies.

Significant global disparities exist in HPV infection burden, with higher rates prevalent in developing and impoverished nations compared to developed countries. Variations in HPV positivity rates and genotype distribution are also observed across different regions within the same country. This study identified an HPV infection rate of 11.31% (64/566) among the eligible participants in Suzhou. This finding is lower than the rates documented in Nanjing (28.79%), highlighting substantial intra-provincial variation (Zhang et al., 2019). These differences may be attributed to regional variations in HPV genotype distribution and the comparatively higher socioeconomic status and health literacy of the Suzhou study population.

Using fluorescent quantitative PCR for genotyping, this study found the most prevalent genotypes to be HPV16, HPV52, and HPV58, each with an infection rate of 1.77%. Single infections were predominant (8.83%), while multiple infections accounted for 2.47%, with HPV35 and HPV52 co-infection being a notable combination. The relationship between multiple HPV infections and cervical cancer risk remains contentious, with studies suggesting either a synergistic effect or competitive inhibition between genotypes, warranting further investigation (Swase et al., 2025).

A significant age-dependent increase in HPV infection was observed (P < 0.001), with the highest rate in women aged ≥50 years (24.37%), followed by those aged 40-49 (8.42%) and ≤39 (7.35%). This trend may be linked to age-related immune decline, hormonal changes, vaginal microenvironment imbalances, and lower health literacy among older women (Yoshikata et al., 2022). Univariate analysis identified a history of secondhand smoke exposure as a significant risk factor (P < 0.05), with an infection rate of 21.57% among exposed women versus 10.29% in non-exposed women.

Smoking is an established independent risk factor for persistent HPV infection, potentially because tobacco metabolites like nicotine disrupt cervical cell cycles and impair local immune responses (Mazarico et al., 2015). Previous study showed that women who continued to smoke more than 10 cigarettes per day for six months were 1.63 times more likely to develop HPV infection than non-smoking women. This phenomenon may be due to the accumulation of harmful substances in tobacco in women’s cervical tissue, resulting in the inability of immune cells to eliminate HPV (Bahmanyar et al., 2012). While some studies corroborate this link, others report no association, indicating a need for larger multi-center studies.

Similarly, a history of underlying diseases and prior cervical surgery were significantly associated with higher HPV infection rates (24.32% vs. 10.40% and 20.75% vs. 10.33%, respectively; P < 0.05). Immunosuppression resulting from underlying conditions increases susceptibility to HPV, while cervical surgery may disrupt the vaginal microenvironment and local immunity, facilitating infection (HALPERT et al., 1986).

HPV vaccination is a cornerstone of global efforts to eliminate cervical cancer. Although Suzhou began offering HPV vaccines in 2017, coverage remains low, with only 14.13% of the study population vaccinated. The nine-valent vaccine covers 47.36% (9/19) of the high-risk genotypes identified in this study. A recent municipal policy providing free HPV vaccination for school-aged girls represents a significant step toward improving coverage and accelerating cervical cancer elimination in China (Zhang et al., 2025). To mitigate HPV risk, targeted health education should be enhanced, particularly for older women, focusing on promoting healthy lifestyles and raising awareness about HPV and cervical cancer. Correcting unhealthy habits can reduce infection rates and ultimately decrease cervical cancer incidence.

This study has several limitations: (1) The data are from a single region and may not be nationally representative; (2) The sample size (n=566) is relatively small, potentially limiting generalizability within Suzhou; (3) The test only covered 21 HPV genotypes, possibly missing infections caused by other genotypes.

In conclusion, this study reveals several key findings regarding HPV infection among women in the Suzhou region. The HPV positive infection rate remains relatively high, with the predominant genotypes identified as HPV16, HPV52, and HPV58. A diverse spectrum of HPV genotypes was observed, with single infections constituting the majority of cases. Women aged 50 years and older demonstrated higher susceptibility to HPV infection. Significant risk factors for genital HPV infection in this population include advanced age, exposure to secondhand smoke, history of underlying medical conditions, and previous cervical surgery. These findings underscore the necessity of tailoring prevention and control strategies to local epidemiology, particularly by aligning vaccination programs with the predominant genotypes (HPV16, 52, and 58) for effective cervical cancer prevention in Suzhou.

## Data Availability

All data produced in the present study are available upon reasonable request to the authors

## Ethical approval

The study was approved by the Human Research Ethics Committees of Suzhou Center for Disease Control and Prevention. All specimens were collected according to the guidelines set by the Suzhou Municipal Hospital Health Management Center. All authors confirm that all methods were performed in accordance with the relevant guidelines and regulations (Declaration of Helsinki).

## Informed Consent Statement

Written informed consents were obtained from the parents of all the enrolled infants before specimen collection.

## Consent for publication

Not applicable

## Authors’ contributions

HC, HL, YZ wrote the main manuscript text and LW, ZZ prepared the figures and tables. All authors reviewed the manuscript.

## Acknowledgements

Not applicable

